# Insights into the practical effectiveness of RT-PCR testing for SARS-CoV-2 from serologic data, a cohort study

**DOI:** 10.1101/2020.09.01.20182469

**Authors:** Zhen Zhang, Qifang Bi, Shisong Fang, Lan Wei, Xin Wang, Jianfan He, Yongsheng Wu, Xiaojian Liu, Wei Gao, Renli Zhang, Wenfeng Gong, Qiru Su, Andrew S Azman, Justin Lessler, Xuan Zou

## Abstract

**Background:** Virologic detection of SARS-CoV-2 through Reverse Transcriptase Polymerase Chain Reaction (RT-PCR) has limitations for surveillance. Serologic tests can be an important complementary approach. We assess the practical performance of RT-PCR based surveillance protocols, and the extent of undetected SARS-CoV-2 transmission in Shenzhen, China.

**Methods:** We followed close-contacts of all PCR-confirmed cases detected since January 2020 and measured anti-SARS-CoV-2 antibodies in PCR-negative contacts 2-15 weeks after initial virological testing by RT-PCR using total Ab ELISA. We assessed rates of undetected infection, performance of RT-PCR over the course of infection, and characteristics of seropositive but PCR-negative individuals.

**Findings:** The adjusted seropositivity rate for total Ab among 880 PCR-negative close-contacts was 4·1% (95%CI 2·8%-5·7%), significantly higher than among residents without known exposure to cases (0·0%, 95%CI 0·0%-1·0%). PCR-positive cases were 8·0 times (RR; 95% CI 5·3-12·7) more likely to report symptoms than the PCR-negative individuals who were seropositive, but otherwise similar. RT-PCR missed 36% (95%CI 28%-44%) of infected close-contacts, and false negative rates appear to be highly dependent on stage of infection.

**Interpretation:** Even rigorous RT-PCR testing protocols may miss a significant proportion of infections, perhaps in part due to difficulties timing testing of asymptomatics for optimal sensitivity. Surveillance and control protocols relying on RT-PCR were, nevertheless, able to contain community spread in Shenzhen.

**Funding:** Bill & Melinda Gates Foundation, Special Foundation of Science and Technology Innovation Strategy of Guangdong Province, and Key Project of Shenzhen Science and Technology Innovation Commission.

## Introduction

Virologic detection of SARS-CoV-2 through Reverse Transcriptase Polymerase Chain Reaction (RT-PCR) is the gold standard for diagnosing infection^1^. Almost all diagnostic testing for COVID-19 is done using PCR-based methods, though rapid antigen tests are quickly gaining in popularity. Like all virologic tests, RT-PCR has imperfect sensitivity^2-4^, and patterns of viral shedding make the chance of testing positive vary over the course of infection^5,6^. Hence, while RT-PCR may be highly accurate at identifying those currently infectious, individuals must be tested at the right time during their infection to be detected, limiting the utility of virologic testing for measuring overall SARS-CoV-2 incidence.

Serologic tests offer an alternative approach for detecting SARS-CoV-2 infection, based on the long lasting immunologic signature it leaves, that can be detected for months, if not years, by measuring circulating antibodies against the virus. In contrast to virologic tests, serologic tests can tell if an individual has been infected long after viral clearance, though these assays also have imperfect sensitivity and specificity^7^.

By utilizing both tests in the same population, we can gain insights into the performance of each. In particular, we can gain understanding of the practical performance of RT-PCR based surveillance if three conditions are met (1) virologic surveillance occurred around the time of potential exposure in a population, (2) the same individuals tested by RT-PCR later received serologic tests, regardless of their virologic test result, and (3) there was a low chance of infection between the periods of virologic and serologic surveillance.

For SARS-CoV-2, Shenzhen, China is one area where all three conditions were met. The Shenzhen Center for Disease Control and Prevention (CDC) implemented extensive contact tracing among local residents during the first wave of the COVID-19 epidemic. During this wave, which started in mid-January, 2020, nearly all infections were among travellers who had been to other parts of China and their close-contacts^8^. After this initial epidemic wave receded, local COVID-19 cases disappeared from the region, and, as of August 1, 2020, all COVID-19 cases have been imported since February 16, 2020 ^9^. During this period, we performed serologic testing among PCR-negative close-contacts of confirmed cases and conducted a serosurvey of local residents without known COVID-19 exposure. Here, we compare the results of these serologic analyses to those of the initial RT-PCR based surveillance to gain insight into the practical performance of RT-PCR based virologic surveillance protocols, and the extent of undetected transmission in the region over this period. Using PCR results from infected individuals, we also characterise the false-positivity rate of RT-PCR over time, both before and after symptom onset.

## Methods

### Study setting, design, and participants

The Shenzhen CDC implemented a surveillance program and strict quarantine policy to monitor travellers from Hubei province, where the first case of COVID-19 was detected, from early January 2020 until the lockdown in Hubei province was lifted around the end of March. Mandatory screening and quarantine of all international travellers started on March 27th. Suspected cases were also detected at local hospitals and through fever screening in neighborhoods.

Contact tracing was used to identify close-contacts of confirmed cases, defined as those who lived in the same apartment, shared a meal, travelled, or socially interacted with an index case from 2 days prior to symptom onset (see Bi *et al*. for a list of 21 symptoms ^8^). Samples were collected from suspected cases and close-contacts using nasopharyngeal swabs, and were tested for SARS-CoV-2 by RT-PCR. By protocol, RT-PCR testing was required for all close contacts at the beginning of quarantine, and release was conditional on two consecutive negative RT-PCR tests from samples collected at least one day apart. Those not contacted within 14 days of the last day of exposure to a case were tested only once. Symptomatic individuals were isolated and treated at designated hospitals regardless of RT-PCR test results for a minimum of 14 days after the last day of putative exposure. All close-contacts, asymptomatic cases testing PCR-positive for SARS-CoV-2, and travellers from Hubei (before lockdown was lifted around end of March) and abroad (after March 27th) were quarantined at centralized facilities, and monitored for 14 days after last day of putative exposure. After the lockdown in Hubei was lifted, domestic travellers were required to test negative by RT-PCR before leaving to come to Shenzhen and upon arrival in Shenzhen.

Between April 12th to May 4th, Shenzhen CDC attempted to recruit all PCR-negative close-contacts of all confirmed COVID-19 cases in Shenzhen for serological testing. Close-contacts were initially contacted by phone. RT-PCR test records before clinical diagnosis were available for all close-contacts, as well as their time and mode of putative exposure. A portion of these PCR-negative close-contacts were included in a previous study of 1,286 close-contacts that characterized epidemiology and transmission parameters of COVID-19 in Shenzhen^8^, hereafter referred to as the Shenzhen cohort. In addition, between April 17th and April 23rd, Shenzhen CDC conducted a serosurvey among 350 volunteers from neighborhoods where no cases were reported in two districts in Shenzhen, Luohu and Longgang, and 50 individuals from neighborhoods in Luohu where cases resided (three PCR-confirmed cases within case neighborhoods, 1·2 per ten thousand). The community sero-survey recruited the same number of volunteers in seven age groups (e.g., 0-9, …, 40-49, 50-59, and 60+ years old).

To estimate sensitivity of RT-PCR tests over the course of infection, we obtained RT-PCR results and time of sampling for a subset of infected individuals. To be included, individuals had to 1) have at least one positive RT-PCR or serologic test, and 2) have been detected through contact tracing, not through symptom-based surveillance. To be included in estimates of sensitivity of RT-PCR based on time before or after symptom onset, cases must have reported symptoms during the initial investigation. For PCR-confirmed cases, PCR-results were available up to the first positive, and potentially one additional confirmatory, RT-PCR test (from a different date).

All close-contacts and local residents gave written informed consent before participating in the serological testing. Contact tracing and RT-PCR testing is part of the continuing public health investigation of an emerging outbreak and therefore the individual informed consent was waived. The study was approved by the ethics committees of Shenzhen CDC. This work was done in support of an ongoing public health response, and hence was determined not to be human subjects research after consultation with the Johns Hopkins Bloomberg School of Public Health institutional review board.

### Laboratory analyses

We assessed anti-SARS-CoV-2 antibodies in participants’ serum using commercially available total Ab, IgG, and IgM ELISA assays that detect antibodies binding SARS-CoV-2 spike protein receptor binding domain (RBD), according to the manufacturer’s instruction (Beijing Wantai Biological Pharmacy Enterprise, Beijing, China; Cat # WS1096). We excluded close-contacts sampled within two weeks of last exposure (n=295) due to the low expected sensitivity of antibody tests during this time window ^10^. Previously published studies reported that the manufacturer’s recommended cutoff for positivity of the total Ab ELISA had sensitivity ranging from 93% (validation cases sampled 7 to 21+ days post symptom onset) to 99% (validation cases sampled 1-43 days post symptom onset) and specificity ranging from 99% to 100% ^11,12^. A combined estimate of sensitivity from the two validation studies was calculated with inverse-variance weights.

We estimated the crude and adjusted sero-positive rate for total Ab, IgG, and IgM among PCR-negative close-contacts in the Shenzhen cohort, other PCR-negative close-contacts outside of the Shenzhen cohort, local residents living in neighborhoods where reported cases resided, and local residents living in neighborhoods where no cases were reported. To account for assay performance, we calculated the adjusted seropositivity rate as *(proportion of positive tests + (spec − 1))/(sens + spec −* 1). We further corrected the seropositivity rate for the neighborhood background infection rate, defined as the adjusted seropositivity rate of local residents living in neighborhoods where reported cases resided^13-15^. We based our analyses on those seropositive for total Ab unless otherwise noted.

### Statistical analyses

We used Poisson regression to estimate the relative risk of developing symptoms comparing the seropositive and seronegative contacts and comparing the PCR-positive contacts and PCR-negative contacts who seroconverted. We used the Breslow Day test to assess heterogeneity in the exposure specific odds-ratio of positivity between serologic and virologic studies^16^. We estimated the number of close-contacts missed by RT-PCR using the adjusted seropositivity rate estimated among all tested PCR-negative close-contacts. Then, using the adjusted seropositivity rate among close-contacts in Shenzhen, we re-estimated the secondary attack rate (SAR) and effective reproductive number (R) in the Shenzhen cohort using methods previously described ^8^. We assumed that the PCR-negative close-contacts with serological test results were representative of all PCR-negative close-contacts in the Shenzhen cohort.

We estimated sensitivity of RT-PCR over time from symptom onset using data from both PCR positive and seropositive symptomatic contacts ^8^. To estimate sensitivity of RT-PCR over time from last exposure to an index case, we also included sero-converted contacts who were asymptomatic. Using an approach similar to that of Kucirka *et* al.^5^, we fitted a Bayesian logistic regression model for test sensitivity with a polynomial spline for time since symptom onset. We assessed the performance of models that incorporated polynomial splines of third to fifth degree with Widely Applicable Information Criterion (WAIC). We implemented this model in the Stan probabilistic programming language and used the *rstan* package to run the model and analyse outputs. We ran 6,000 iterations (four chains of 1500 iterations each with 250 warm-up iterations) and assessed convergence visually and using the R-hat statistic. All reported estimates are means of the posterior samples with the 2·5th and 97·5th percentiles of this distribution reported as the 95% confidence interval (CI). From the expected sensitivity of RT-PCR, we calculated the expected false negative rate on each day (1-sensitivity). We estimated sensitivity of RT-PCR up to a week after symptom onset because RT-PCR results after clinical diagnosis (which usually happened a few days after symptom onset and requires 1) a positive PCR result and 2) symptom onset or chest CT abnormality) were generally not available. We also fit generalized additive models (GAM) for test sensitivity as a function of the time from symptom onset (and time from last exposure to an index case) using a thin plate regression spline^17^ as implemented in the mgcv package in R to data form Shenzhen alone and a pooled dataset including data from Kucirka *et al.^5^*.

## Results

As of August 1, 2020, 348 imported cases from other parts of China, 39 cases from abroad, and 75 locally-transmitted cases have been detected in Shenzhen (cumulative incidence 0·35 per 10,000). The majority of the locally-transmitted cases (63/75) were close-contacts of a confirmed imported case. At the time of writing, the vast majority of cases in Shenzhen (417/462), including locally-transmitted and imported cases, were confirmed during the first wave of the outbreak that ended on February 21, 2020.

From April 12 to May 4, 2020, Shenzhen CDC successfully contacted and collected serological samples from 2,345 of 4,422 PCR-negative close-contacts of COVID-19 cases. Among these, 1,175 were contacts of cases diagnosed in Shenzhen that had contact tracing records. Sera from 880 of these contacts were collected more than two weeks after exposure to an index case (Figure 1). Among the 880 close-contacts, the average age was 34·1 years (IQR=24·0, 44·0), and the majority were females (52·3%, n=460; Appendix Table 1). PCR-negative close-contacts had, on average, 32 RT-PCR tests each (IQR=2·0, 3·0; range=1,10) before the end of quarantine, and 814% (n=716) of them had more than one RT-PCR test (Figure 2). Average time of serological testing from last exposure to an index case was 71·4 days (IQR=56·0, 87·0) (Figure 1). Among these close-contacts, 27·6% (n=243) reported “frequent” contact with the index case, 28·4% (n=250) reported “moderate” contact and 44·0% (n=387) reported “rare” contact.

**Figure 1.**
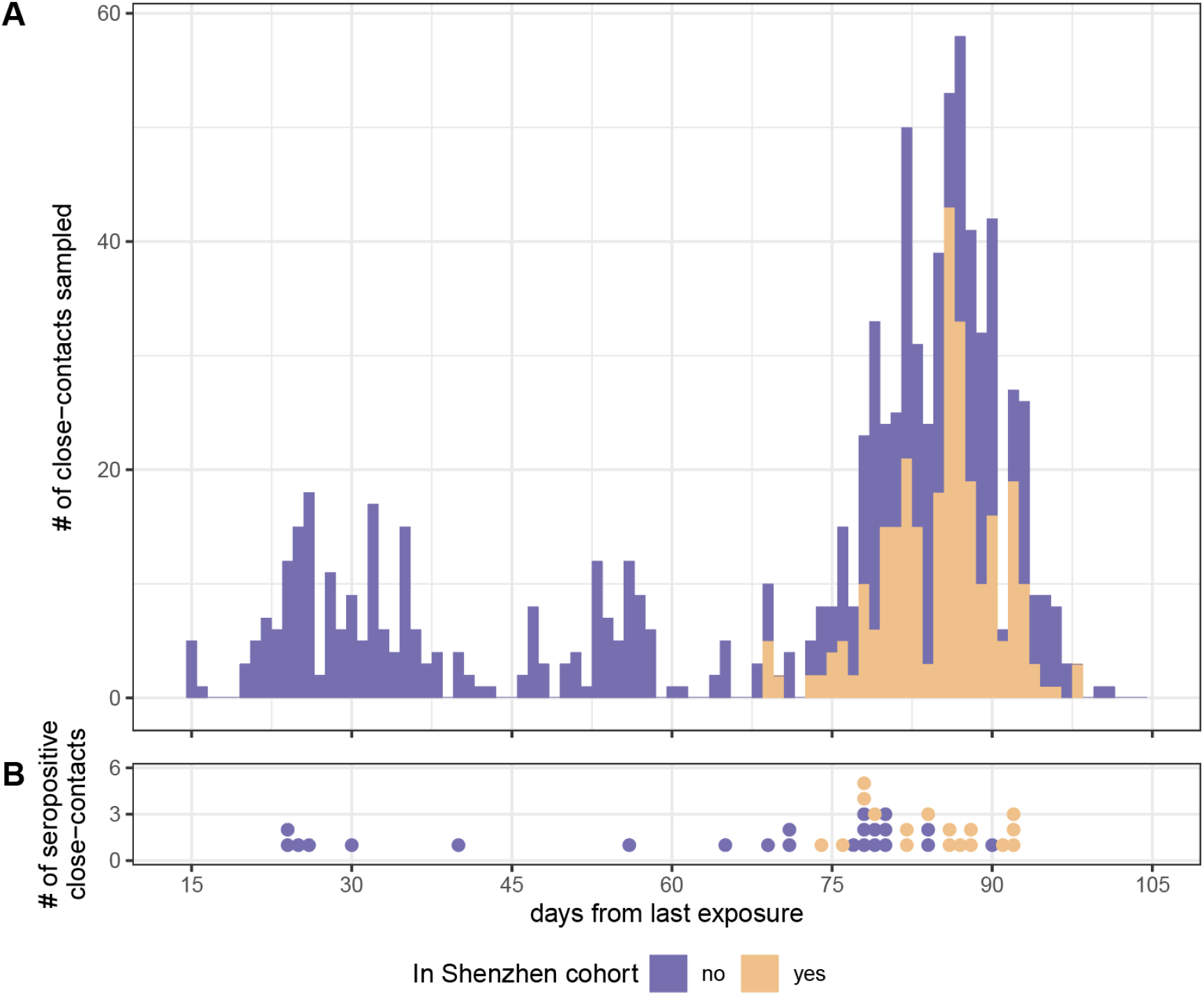
Time of serological testing from last putative exposure to an index case (A) among all PCR-negative close-contacts and (B) among those tested positive. Those in and out of the Shenzhen cohort were colored in purple and orange. All close-contacts had one serological test per person.

Forty of the 880 PCR-negative close-contacts (4·5%) were positive for total Ab antibodies; of these, 34 (3·9%) tested positive for IgG and 16 (1·8%) for IgM. Adjusting for assay performance ^11,12^, we estimated a seropositivity rate of 4·0% (95%, CI 2·8% to 5·6%, Table 2). The seropositivity rate among PCR-negative close-contacts in Shenzhen was significantly higher than local residents without known exposure to cases; only one person (0·29%, 1/350) was seropositive in neighborhoods where no cases resided and no one (0/50) was seropositive in neighborhoods where reported cases resided.

Among PCR-negative close-contacts sampled within 60 days from the last day of exposure to a known case, the unadjusted seropositivity rate was 2·9% (7/241), it was 5·6% (29/546) for those sampled 61-90 days after, and 4·3% (4/93) for over 90 days (p-value=0·33; Table 1). The overall proportion seropositive among PCR-negative close-contacts did not differ significantly by age or sex (Table 1).

**Table 1.** Characteristics of PCR-negative close-contacts in Shenzhen who tested positive and negative for total Ab ELISA.

|  | Seronegative close-contacts (N=840) | Seropositive close-contacts (N=40) | P-value |
| --- | --- | --- | --- |
| **Sex** |  |  |  |
| Female | 52.1% (438) | 55.0% (22) | 0.85 |
| Male | 47.9% (402) | 45.0% (18) | - |
| **Age** |  |  |  |
| 0-9 years | 8.9% (75) | 12.5% (5) | 0.73 |
| 10-19 years | 7.9% (66) | 5.0% (2) | - |
| 20-29 years | 22.6% (190) | 25.0% (10) | - |
| 30-39 years | 26.4% (222) | 27.5% (11) | - |
| 40-49 years | 18.0% (151) | 7.5% (3) | - |
| 50-59 years | 9.4% (79) | 12.5% (5) | - |
| 60-69 years | 5.1% (43) | 7.5% (3) | - |
| >=70 years | 1.7% (14) | 2.5% (1) | - |
| **Symptomatic** |  |  |  |
| No | 99.0% (832) | 92.5% (37) | 0.0036 |
| Yes | 1.0% (8) | 7.5% (3) | - |
| **Contact Frequency** |  |  |  |
| Rare | 45.5% (382) | 12.5% (5) | <0.0001 |
| Moderate | 29.4% (247) | 7.5% (3) | - |
| Often | 25.1% (211) | 80.0% (32) | - |
| **# of RT-PCR tests before the end of quarantine** | 3.1 (2,3) | 3.9 (2, 5.3) | 0.070 |
| <=2 tests | 68.9% (579) | 52.5% (21) | 0.049 |
| >2 tests | 31.1% (261) | 47.5% (19) | - |
| **Days from last exposure to a case to serologic testing** |  |  |  |
| <=60 | 27.9% (234) | 17.5% (7) | 0.33 |
| 61-90 | 61.5% (517) | 72.5% (29) | - |
| >90 | 10.6% (89) | 10.0% (4) | - |

**Table 2.** Crude and adjusted seropositivity rate of SARS-CoV-2 antibody by total Ab, IgG, and IgM ELISA among individuals with and without exposure to cases diagnosed with COVID-19 in Shenzhen.

|  | Crude estimates |  |  | Adjusted estimates<sup>†</sup> |  |  |
| --- | --- | --- | --- | --- | --- | --- |
|  | % positive for total Ab (95%CI; # seropositive) | % positive for IgG (95%CI; # seropositive) | % positive for IgM (95%CI; # seropositive) | % positive for total Ab (95% CI) | % positive for IgG (95%CI) | % positive for IgM (95%CI) |
| PCR-negative close-contacts (N=880) | 4.5<br>(3.4, 6.1; n=40) | 3.9<br>(2.8, 5.4; n=34) | 1.8<br>(1.1, 2.9; n=16) | 4.1<br>(2.9, 5.7) | 1.7<br>(0, 5.3) | 0.55<br>(0, 1.8) |
| PCR-negative close-contacts in the Shenzhen cohort (N=288) | 5.9<br>(3.7, 9.2; n=17) | 5.6<br>(3.4, 8.8; n=16) | 3.1<br>(1.7, 5.8; n=9) | 5.5<br>(3.2, 8.9) | 3.5<br>(0, 8.7) | 2.0<br>(0.37, 5.1) |
| Neighborhood without cases (N=350) | 0.29<br>(0, 1.6; n=1) | 0<br>(0, 1.1; n=0) | 0<br>(0,1.1; n=0) | 0<br>(0, 1.1) | 0<br>(0, 0.97) | 0<br>(0,0) |
| Neighborhood with cases (N=50) | 0<br>(0, 1.1; n=0) | 0<br>(0,1.1; n=0) | 0<br>(0, 1.1; n=0) | 0<br>(0, 6.8) | 0<br>(0, 7.0) | 0<br>(0, 6.6) |

<sup>†</sup> Sensitivity and specificity of total Ab used for calculating adjusted estimates of seropositivity rates were 98% and 99%, 96% and 98% for IgG, and 90% and 99% for IgM <sup>11</sup>. Sensitivity and specificity were based on results from GeurtsvanKessel et al. and Lassaunière et al.. Sensitivity and specificity of the IgG ELISA were based on an assay validation study conducted by Shenzhen CDC (unpublished result). Validation samples were collected >14 days after symptom onset in 23 hospitalized COVID-19 cases and compared to 44 pre-pandemic controls.

Only three of the 40 (7·5%) seropositive contacts who were PCR-negative reported symptoms between the last date of putative exposure and end of quarantine (all three had fever, two had signs of lung infection, and one had nausea and headache) (Figure 2, Table 1). Nevertheless, seropositive but PCR-negative contacts were more likely to report symptoms than those who were both PCR-negative and seronegative (RR = 7·9; 95%CI, 1·7 to 27·2; p-value = 0·0023). In the Shenzhen cohort, PCR-positive cases were 8·0 times (RR, 95% CI, 5·3 to 12·7, p-value <0·0001) more likely to report symptoms than the PCR-negative individuals who seroconverted (366/391 vs. 2/17). In this PCR-negative group, there was no significant difference in the number of PCR-tests seropositive and seronegative contacts received (seropositives had on average 0·81 more tests, 95% CI, -0·041 to 1·7).

**Figure 2.**
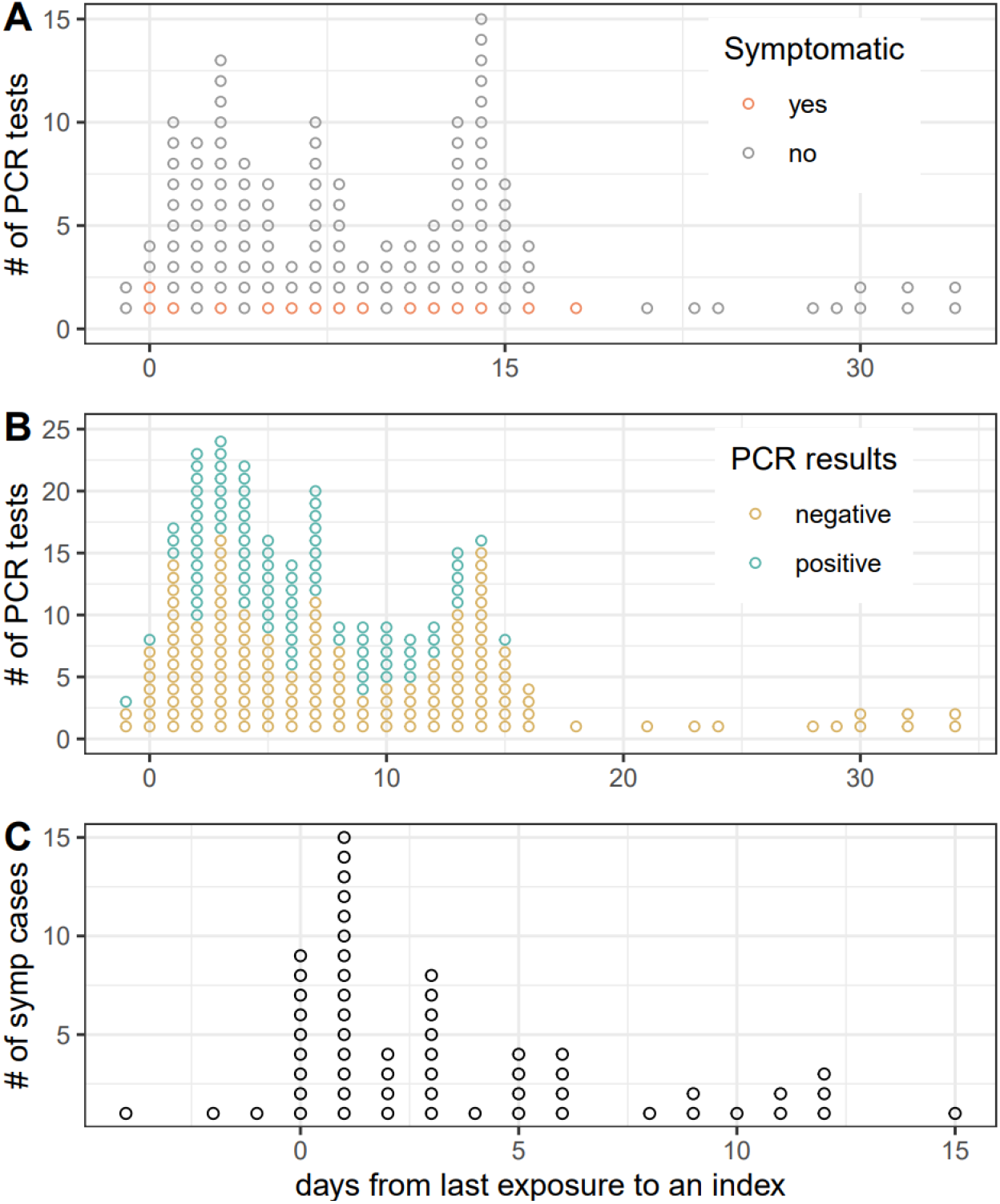
Time of RT-PCR test and time of symptom onset from last exposure to an index case. A) Time of RTPCR tests among seropositive close-contacts who only had PCR-negative test (N=40). Those who ever showed (n=3) and never showed symptoms (n=37) before the end of quarantine were colored in red and grey respectively. B) Time of RT-PCR tests among infected contacts who were either diagnosed with COVID-19 by RT-PCR (n=75) or had only PCR-negative test but later tested seropositive (n=40). Positive and negative PCR results were colored in green and yellow respectively. C) Time of symptom onset from last exposure to an index case among symptomatic and infected contacts who were either diagnosed with COVID-19 by RT-PCR (n=57) or had only PCR-negative test but later tested seropositive (n=3).

Using the adjusted seroprevalence of 4·1% (95%CI, 2·9% to 5·7%) among PCR-negative close-contacts (Table 2), we estimated that RT-PCR missed 36% (95%CI, 28% to 44%; 48 infections) of all infected close-contacts in the Shenzhen cohort. Adjusting for these missed infections increases our estimate of the effective reproductive number by 34% to 0·56 (95%CI, 0·45 to 0·67). Overall, our estimates of the secondary attack rate and household secondary attack rates increased to 108% and 159%, compared with previous estimates of 6·6% and 11·2% respectively.

Among the 288 members of the previously published Shenzhen cohort that were included in this serosurvey, we compared risk factors for seropositivity in PCR-negatives with the originally published results (Figure 1, Appendix Table 2). As noted above, those testing PCR-positive were eight times as likely to report symptoms than their PCR-negative, seropositive counterparts. However, in the Shenzhen cohort there was no significant difference in the odds-ratio of testing positive for any exposure between the serologic and virologic studies.

We examined the variability in the false negative rate of PCR-based testing over the course of infection using RT - PCR results from 60 symptomatic cases who were either diagnosed with COVID-19 by RT-PCR (n=57) or had only PCR-negative test but later tested seropositive (n=3) (Figure 3a). Seven cases had their first RT-PCR test before symptom onset and 20 on the day of symptom onset. Based on the best fit polynomial spline model (three degrees of freedom, Appendix Table 3) we estimate the probability of having a false negative results, to be 34% (95% CI, 21% to 51%) on the day of symptom onset, decreasing to a low of 11% (95% CI, 5% to 21%) four days after symptom onset. Uncertainty in the false negative rate is high in the days prior to symptom onset, but we estimate a smooth increase in the probability of a false negative rate as we move earlier in the course of infection, reaching 100% at 5 days prior to symptom onset; but 95% confidence intervals include false negative rates lower than 75% even 8 days prior to symptom onset.

**Figure 3.**
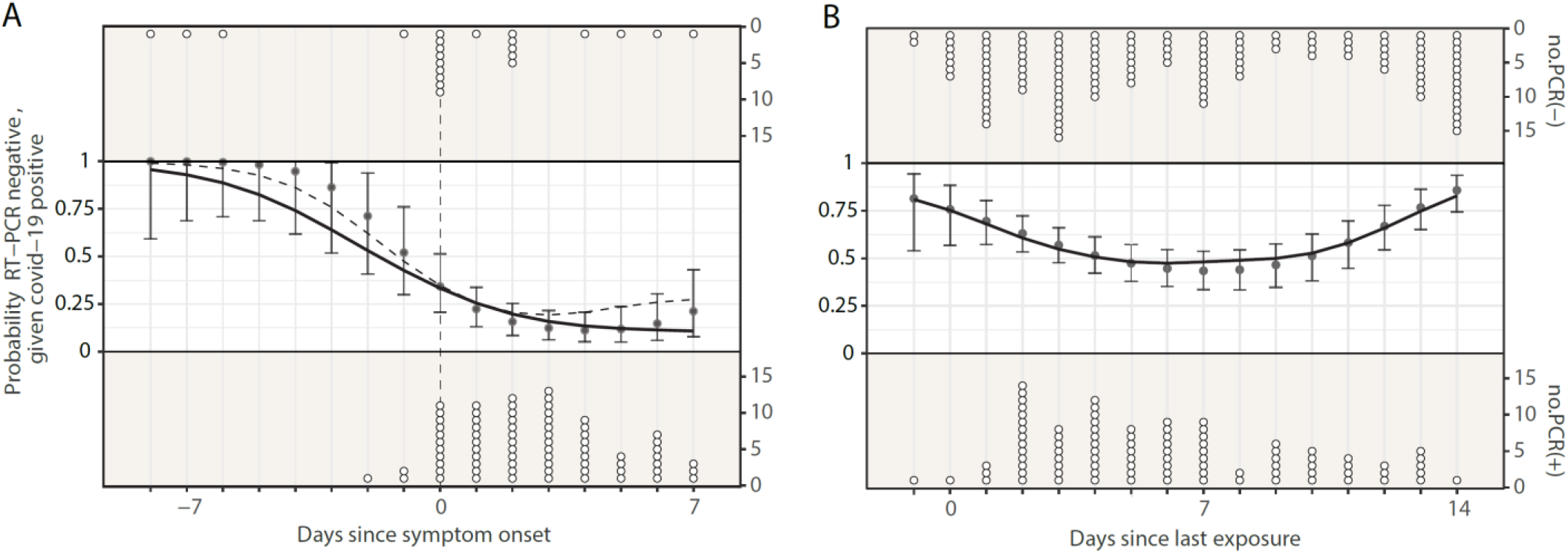
False negative rate of RT-PCR of nasopharyngeal swab by A) time since symptom onset and B) time since last exposure to an index case. Point estimates and confidence intervals represent estimates from the Bayesian logistic regression model for test sensitivity with a polynomial spline of third degree. Solid curve represents estimates from the GAM model fitted to Shenzhen data only. Dashed curve represents estimates from the GAM model fitted to the combined Shenzhen data and pooled data from Kucirka *et* al.^5^. Dots on top and bottom of the figures indicate timing of negative and positive PCR results respectively. Vertical dashed line corresponds to time of symptom onset.

## Discussion

The specific circumstances of the initial SARS-CoV-2 epidemic in Shenzhen, China allowed us to use serologic data to gain additional insight into the epidemiology of the virus and the practical performance of RT-PCR as a surveillance tool. We find that 4·5% of PCR-negative close-contacts in Shenzhen were seropositive for anti-SARS-CoV-2 antibodies. Thus, despite a rigorous testing regime, virologic testing failed to detect 30-40% of infections. Even with these limitations, the serologic evidence suggests that the overall control program, which relied on RT-PCR to clear people from isolation or quarantine, was successful in containing the virus; as the rate of seropositivity outside of close-contacts was virtually zero. Those testing PCR-negative but seropositive were significantly less likely to have symptoms than those testing PCR-positive, potentially reflecting difficulties in appropriately timing virologic testing when there is no outward indication of the timing of viral shedding. Consistent with this latter point, we found considerable variation in the chance of an infected individual testing negative over the course of their infection.

It may be that RT-PCR test results are correlated with transmissibility, but we were unable to assess that in this study. Strict quarantine practices were in place, requiring all close-contacts to be quarantined in centralized facilities for about 2 weeks after exposure. Critically, individuals remained in quarantine regardless of symptoms or test results. The vast majority (92·5%) of individuals who were seropositive, but PCR-negative, reported no symptoms. While this may stem from correlations between viral shedding and development of symptoms, it also may highlight a challenge in using virologic testing for asymptomatic surveillance. The period in which RT-PCR testing is highly sensitive is relatively short^5,6^, and sensitivity peaks around the time of, or shortly after, symptom onset. Hence, without symptoms as an indicator of when to test, it may be difficult to capture patients in this period. This phenomena is not unique to SARS-CoV-2, and studies of influenza and other acute respiratory viruses have shown serologic attack rates 2-3 times virological attack rates^18,19^. Therefore, while RT-PCR is an invaluable diagnostic tool, it has important limitations as a tool for surveillance or an outcome in risk factor studies.

The study has important limitations. We were unable to obtain serologic data on PCR-positive cases, so we could not estimate the practical sensitivity of the serologic tests. Sample size was limited, particularly for estimating risk factors in the Shenzhen cohort, and only 20% of PCR-negative contacts both could be recontacted and had detailed data from the initial contact tracing. The serum samples were collected between two weeks to four months after the last day of exposure to a known case. Although we expect most cases to seroconvert in the first month after exposure, time to sampling may not have been enough for some close-contacts to seroconvert. While the serologic tests have imperfect specificity, seroprevalence among PCR-negative close-contacts were much higher than the seroprevalence among those without known exposure to cases, lending confidence that the observed undetected infections are real. Seropositivity rate among the subset in the Shenzhen cohort is slightly higher than the rate among all close-contacts in Shenzhen (5·4% vs. 4·0%), possibly due to the more frequent self-reported exposure between index cases and contacts in the cohort. The extent of undetected transmission estimated using the lower seropositivity rate in Shenzhen may provide an underestimate of the true extent.

We cannot completely rule out the possibility that secondary exposure to cases other than their index cases led to seroconversion in close-contacts because we did not ascertain recent travel history of these individuals. However, the epicenter of COVID-19 in China was under lockdown from January 23 to March 25th and mandatory RT-PCR testing of all international travellers arriving in Shenzhen was enforced starting March 27th. As a result, the chance of these individuals travelling to high risk areas before serological screening that were performed in April was low. Given the low seropositivity rate in community samples without direct exposure to a known case, we believe seroconversion among close-contacts was most likely due to exposure to the index case. We cannot rule out infection by the index case after initial quarantine, as there have been reports of viral shedding after multiple PCR-negative tests^20^.

Overall, this study gives important insights into the practical limitations of RT-PCR based virologic surveillance for SARS-CoV-2. While virologic testing should remain the bedrock of disease control programs and patient diagnostics, it, like all tools, is imperfect. Hence, it is essential that we weigh all the evidence when making clinical and public health decisions, and not rely on test results alone. While serologic testing is an important supplement to RT-PCR for broad scale surveillance and scientific study, it is a fundamentally different tool and cannot replace virologic testing. Innovations to both improve accuracy of virologic testing, and allow for more frequent less invasive testing are, therefore, sorely needed. Still, the Shenzhen experience highlights that even with the imperfect tools we have available, SARS-CoV-2 control is possible through rigorous and aggressive surveillance, isolation and quarantine combined with broader public health action.

## Data Availability

Summary data are available upon reasonable request

## Acknowledgement

We thank all patients, close contacts, and their families involved in the study, as well as the front-line medical staff and public health workers who collected these important data. The study was funded by Bill & Melinda Gates Foundation (INV-006376), Special Foundation of Science and Technology Innovation Strategy of Guangdong Province of China (N0.2020B1111340077), and Key Project of Shenzhen Science and Technology Innovation Commission, Shenzhen, China (202002073000003).

## >Supplemental material

**Appendix Table 1.** Characteristics of PCR-negative close-contacts in the serological survey.

|  | All PCR-negative close-contacts in the serosurvey (N=880) | PCR-negative close-contacts in the Shenzhen cohort (N=288) | PCR-negative close contacts not in the Shenzhen cohort (N=592) | P-value |
| --- | --- | --- | --- | --- |
| **Sex** |  |  |  |  |
| Female | 52.3% (460) | 54.2% (156) | 51.4% (304) | 0.48 |
| Male | 47.7% (420) | 45.8% (132) | 48.6% (288) | - |
| **Age** |  |  |  |  |
| 0-9 years | 9.1% (80) | 12.5% (36) | 7.4% (44) | 0.0047 |
| 10-19 years | 7.7% (68) | 7.6% (22) | 7.8% (46) | - |
| 20-29 years | 22.7% (200) | 17.7% (51) | 25.2% (149) | - |
| 30-39 years | 26.5% (233) | 26.0% (75) | 26.7% (158) | - |
| 40-49 years | 17.5% (154) | 16.7% (48) | 17.9% (106) | - |
| 50-59 years | 9.5% (84) | 9.4% (27) | 9.6% (57) | - |
| 60-69 years | 5.2% (46) | 8.7% (25) | 3.5% (21) | - |
| >=70 years | 1.7% (15) | 1.4% (4) | 1.9% (11) | - |
| **Symptomatic** |  |  |  |  |
| No | 98.7% (869) | 97.2% (280) | 99.5% (589) | 0.012 |
| Yes | 1.3% (11) | 2.8% (8) | 0.51% (3) | - |
| **Contact Frequency** |  |  |  |  |
| Rare | 44.0% (387) | 16.3% (47) | 57.4% (340) | <0.0001 |
| Moderate | 28.4% (250) | 36.1% (104) | 24.7% (146) | - |
| Often | 27.6% (243) | 47.6% (137) | 17.9% (106) | - |
| **# of RT-PCR tests before the end of quarantine** | 3.2 (IQR=2,3) | 2.1 (IQR=2,2) | 3.7 (IQR=2,4) | <0.0001 |
| <=2 tests | 68.2% (600) | 87.5% (252) | 58.8% (348) | <0.0001 |
| >2 tests | 31.8% (280) | 12.5% (36) | 41.2% (244) | - |
| **Days from last exposure to a case to serologic testing** | 71.4 (IQR=56,87) | 85.1 (IQR=82,88) | 64.7 (IQR=36,86) | <0.0001 |
| <=60 | 27.4% (241) | 0% (0) | 40.7% (241) | <0.0001 |
| 61-90 | 62.0% (546) | 85.4% (246) | 50.7% (300) | - |
| >90 | 10.6% (93) | 14.5% (42) | 8.6% (51) | - |

**Appendix Table 2.** Comparison of exposure specific positivity rates in serological and virological testing, and test for heterogeneity in the exposure specific odds-ratio of positivity between serologic and virologic studies^16^.

|  | # PCR-negative contacts with serologic testing | # Sero-positive | Seropositivity rate among PCR-negative contacts (95%CI) | Number of contacts† | Number of PCR-confirmed contacts† | P-value |
| --- | --- | --- | --- | --- | --- | --- |
| **Sex** |  |  |  |  |  |  |
| Female | 156 | 9 | 5.8% (3.1, 10.5) | 558 | 58 | 0.16 |
| Male | 132 | 8 | 6.1% (3.1, 11.5) | 486 | 26 | ref |
| **Age** |  |  |  |  |  |  |
| 0-9 years | 36 | 2 | 5.6% (1.5, 18.1) | 148 | 11 | 0.94 |
| 10-19 years | 22 | 0 | 0 | 85 | 6 | 0.27 |
| 20-29 years | 51 | 4 | 7.8% (3.1, 18.5) | 114 | 7 | 0.64 |
| 30-39 years | 75 | 7 | 9.3% (4.6, 18.0) | 268 | 16 | 0.45 |
| 40-49 years | 48 | 0 | 0 | 143 | 7 | 0.19 |
| 50-59 years | 27 | 2 | 7.4% (2.1, 23.4) | 110 | 10 | ref |
| 60-69 years | 25 | 2 | 8.0% (2.2, 25.0) | 130 | 20 | 0.64 |
| >=70 years | 4 | 0 | 0 | 72 | 7 | 0.56 |
| **Contact Frequency** |  |  |  |  |  |  |
| Rare | 47 | 0 | 0 | 230 | 1 | 0.72 |
| Moderate | 104 | 2 | 1.9% (0.53, 6.7) | 305 | 9 | ref |
| Often | 137 | 15 | 10.9% (6.7, 17.3) | 555 | 71 | 0.76 |
| **Contact type: household** |  |  |  |  |  |  |
| Yes | 143 | 13 | 9.1% (5.4, 14.9) | 686 | 77 | 0.11 |
| No | 124 | 3 | 2.4% (0.83, 6.9) | 456 | 4 | ref |
| **Contact type: travel** |  |  |  |  |  |  |
| Yes | 54 | 4 | 7.4% (2.9, 17.6) | 318 | 18 | 0.35 |
| No | 213 | 12 | 5.6% (3.3, 9.6) | 824 | 63 | ref |
| **Contact type: meal** |  |  |  |  |  |  |
| Yes | 153 | 12 | 7.8% (4.5, 13.2) | 707 | 61 | 0.78 |
| No | 114 | 4 | 3.5% (1.4, 8.7) | 435 | 20 | ref |

† Results from the 5th and 6th columns were reported in Table 3 of Bi *et al.*<sup>8</sup>.

**Appendix Table 3.** Performance of polynomial spline models assessed with Widely Applicable Information Criterion (WAIC)

| Degree of freedom of polynomial spline | Models for estimating test sensitivity from symptom onset | Models for estimating test sensitivity from last day of exposure to a case | Sensitivity analysis: models for estimating test sensitivity from symptom onset, excluding test results after the first positive test |
| --- | --- | --- | --- |
| 3rd | 43.5 | 72.2 | 37.3 |
| 4th | 47.9 | 72.8 | 41.6 |
| 5th | 50.8 | 74.7 | 44.3 |

